# Efficacy of Acceptance and Commitment Therapy in Daily Life (ACT-DL) in early psychosis: Results from the multi-center INTERACT randomized controlled trial

**DOI:** 10.1101/2021.05.28.21257986

**Authors:** Inez Myin-Germeys, Evelyne van Aubel, Thomas Vaessen, Henrietta Steinhart, Annelie Klippel, Ginette Lafit, Wolfgang Viechtbauer, Tim Batink, Ruud van Winkel, Mark van der Gaag, Therese van Amelsvoort, Machteld Marcelis, Frederike Schirmbeck, Lieuwe de Haan, Ulrich Reininghaus

## Abstract

**Importance:** Treatment in the early stages of psychosis is crucial to prevent poor clinical and social outcomes. Currently, no preventive interventions are available that reduce psychotic distress, or affective and negative symptoms as well as functioning, calling for more and dedicated treatments for these.

**Objective:** To investigate the efficacy of Acceptance and Commitment Therapy in Daily Life (ACT-DL), combining face-to-face therapy with an Ecological Momentary Intervention (EMI), in addition to treatment as usual for psychotic distress, in comparison to treatment as usual only.

**Design:** This single-blinded randomized clinical INTERACT trial investigated participants post-intervention and at 6 and 12-month follow-up. Participants were recruited between June 1, 2015 and December 31, 2018. Assessors were blinded to treatment allocation.

**Setting:** INTERACT is a multi-center trial recruiting participants from secondary mental health services in 5 regions in Belgium and The Netherlands.

**Participants:** The sample was a referral sample of individuals aged 15-65 years with a clinically established UHR or FEP status.

**Interventions:** Individuals were randomly assigned (1:1) to ACT-DL, consisting of 8 ACT sessions augmented with an EMI app in addition to treatment as usual, or to treatment as usual only.

**Main outcomes and measures:** The primary outcome was a reduction in psychotic distress as assessed with CAARMS at post-intervention, 6-and 12-month follow-up. Secondary outcomes included symptom severity (measured with BPRS and BNNS), functioning (measured with SOFAS and SFS) and momentary psychotic distress (measured with the Experience Sampling Method, a structured diary technique). All analyses were described in the trial protocol and in a postregistration on the open-science framework, prior to accessing the data.

**Results:** Of the 196 individuals assessed for eligibility, 148 were randomized to ACT-DL+TAU (n=71) or TAU (n=77) (72 female (49%), average age 25 (SD = 6), 71 FEP (48%)). 115 (78%) provided primary outcome data at least at one follow-up assessment. There was no evidence of a greater reduction in CAARMS distress in ACT-DL+TAU compared to TAU (χ^2^(3)=2.38; p=.50). However, general psychopathology (χ^2^(3)=14.44; p=.002); affective (χ^2^(3)=8.55; p=.04) and negative symptom severity (χ^2^(3)=19.96; p<.001) as measured with the BPRS was reduced, as well as negative symptoms as assessed with BNNS (χ^2^(3)=15.96; p=.001) in. Furthermore, global functioning improved (χ^2^(3)=8.72; p=.033) in ACT-DL+TAU compared to TAU, whereas social functioning failed to reach significance (χ^2^(3)=7.41; p=.060). Finally, a clear and significant reduction was found in momentary psychotic distress (χ^2^(3)=21.56; p<0.001), whereas no effects were found for momentary psychotic experiences (χ^2^(3)=1.02; p=.599), momentary positive (χ^2^(3)=4.17; p=.124) or negative (χ^2^(3)=2.78; p=.249) affect. No serious adverse events directly related to the therapy occurred.

**Conclusions and relevance:** INTERACT did not support a significant effect on psychotic distress as assessed with the CAARMS. However, significant improvements were found for momentary psychotic distress, global functioning and negative symptomatology. These results are promising given that these latter problems are among the hardest to treat.

**Trial Registration:** Dutch Trial Register: NTR4252

**Key Points:** *Question:* Can Acceptance and Commitment Therapy (ACT) in Daily Life, a combined face-to-face ACT intervention with a digital daily life Ecological Momentary Intervention, reduce psychotic distress in the early stages of psychosis?

*Findings:* This randomized clinical trial of 148 individuals in the early stages of psychosis (UHR or FEP) found no evidence that ACT-DL improved psychotic distress at post-intervention, 6 or 12-month follow-up over and above treatment as usual. However, significant effects were found for momentary psychotic distress, negative symptoms and functioning.

*Meaning:* Whereas the blended care approach of face-to-face ACT with the ACT-DL EMI did not improve psychotic distress over and above treatment as usual, it provided a promising avenue for the treatment of momentary psychotic distress as well as for negative symptoms and improving functioning, some of the hardest to treat problems in individuals with UHR and FEP.

## Introduction

Treatment in the early stages of psychosis, including both Ultra-High-Risk (UHR) and First-Episode Psychosis (FEP)^1–5^, is crucial to prevent transition to more severe stages of illness. While most studies have targeted transition to psychotic disorder or reduction of positive symptoms as their main outcome, distress associated with psychotic symptoms has been identified as a major driver of illness^6^. Also, when investigated at a momentary level using Experience Sampling Methodology (ESM), psychotic distress is prominently present in the early stages of psychosis^7–9^. Furthermore, treatment should target a wider range of symptoms including negative and affective symptoms, while also improving global and social functioning^10^.

A comprehensive meta-analysis of preventive interventions for individuals at ultra-high risk for psychosis found no effects of either psychological or pharmacological interventions on psychotic distress, nor on affective symptoms, negative symptoms and functioning^11^, calling for more and dedicated treatments for these. Acceptance and Commitment Therapy (ACT), a next-generation Cognitive Behavioral Therapy aimed at enhancing individuals’ psychological flexibility, may help patients to handle distress^12^. While ACT components targeting acceptance are likely to be effective in attenuating distress, ACT components targeting commitment may enhance reward-related motivated action. There is good evidence on the feasibility and acceptability of ACT in people with psychosis^13,14^. However, meta-analytic evidence on a small number of studies found limited effects in individuals with an established psychotic disorder^15^. We therefore developed ACT in Daily Life (ACT-DL) for enhancing the therapeutic effects of ACT under real-world conditions^16–19^. ACT-DL builds on the principles of Ecological Momentary Interventions (EMI)^17,20^, providing treatment in real-time. While pilot studies provided evidence on acceptability and feasibility of ACT-DL in both a clinical sample of patients with mental health disorders^21^, as well as in emerging adults with subthreshold levels of depression and psychosis^22^, robust, trial-based evidence on its effects in the early stages of psychosis is lacking.

The aim of the current INTERACT study was to test the efficacy of ACT-DL on reducing psychotic distress (primary outcome) as well as on reducing intensity of psychotic symptoms, general psychopathology and negative symptoms and improving global and social functioning (secondary outcomes) and on improving momentary psychotic distress, psychotic experiences and mood (secondary outcomes measured with the ESM) at post-intervention and 6-and 12-month (for non-ESM outcomes only) follow-ups in patients with UHR and FEP. In an a priori planned subgroup analysis, the effects of ACT-DL on the primary outcome in UHR compared with FEP individuals was investigated. In a more exploratory a priori planned sensitivity analysis, we investigated whether the reduction in psychotic distress is greater for TAU including Cognitive Behaviour Therapy for psychosis (CBTp) compared to TAU as well as for ACT-DL+TAU compared to CBTp+TAU.

## Methods

### Study design

This multi-center single-blind randomized controlled trial (Dutch Trial Register: NTR4252) tested the efficacy of adding ACT-DL to treatment as usual (TAU) compared to TAU in reducing psychotic distress in individuals with UHR or FEP. The trial was conducted at secondary mental health services in five regions in the Netherlands and Belgium. Recruitment took place between June 1, 2015 and December 31, 2018. The study received ethical approval from the MERC at Maastricht University Medical Centre, the Netherlands (reference: NL46439.068.13) and the University Clinic Leuven, Belgium (reference: B322201629214). The trial protocol is published^23^ and the study was postregistered on the open science framework^1^ (see eMethods 1). Mediation analyses as well as the investigation of acceptability and treatment adherence described in the protocol will be reported separately. This study followed the Consolidated Standards of Reporting Trials (CONSORT) reporting guideline.

### Participants

Individuals were referred to the study by their treating clinician. Inclusion criteria were: age 15-65 years; meeting criteria for UHR (without prior use of antipsychotic medication for psychotic symptoms) or FEP (onset within last 3 years) as assessed by the Comprehensive Assessment of At Risk Mental State (CAARMS)^2^ and Nottingham Onset Schedule^24^; sufficient command of the Dutch language; and ability to provide written informed consent. Exclusion criteria included a primary diagnosis of alcohol/substance abuse or dependence, assessed with the Mini-International Neuropsychiatric Interview^25^; and severe endocrine, cardiovascular or brain disease.

### Randomization and masking

Participants were randomized (1:1) after baseline assessment through a computer-generated sequence. Block randomization was carried out in blocks of six participants, with stratification for the five regions and for group (UHR and FEP, expecting a 50:50 ratio). Trained research assessors were blind to allocation. Any breaks in blinding were documented and another researcher was allocated to complete assessments.

### Interventions

Participants allocated to TAU received standard care delivered according to national service guidelines and protocols by their responsible clinician. Standard mental health care included manualized CBTp at some sites. Individuals in the ACT-DL condition received TAU with the exception of manualized CBTp.

ACT-DL consisted of eight manualized ACT sessions (45-60 minutes), administered face-to-face by a trained clinician and an ACT-DL EMI to apply the learned skills in their daily lives. After a psycho-education session, patients received six ACT sessions based on a modified version of ACT for people with psychosis^12,13,26–28^, in the final session all six components were integrated. The ACT-DL EMI prompted participants at eight semi-random moments per day for three days after each session (starting from session two), with a brief questionnaire on their current mood, psychotic experiences and activities, as well as providing an exercise or metaphor of the ACT component covered in the previous session. In addition, participants could do ACT exercises at moments when they were most needed. After completion of the intervention period, participants did no longer have access to the app (see our study protocol^23^ and Vaessen et al., 2019^16^ for more information). Treatment fidelity was rated based on a random selection of audiotapes of three training sessions per participant (range 0-12.6)^23^.

### Measurements

Assessments were conducted before randomization (‘baseline’), after the 8-week intervention period (‘post-intervention’), and after 6-month (all outcomes) and 12-month (all non-ESM outcomes) follow-up.

### Outcomes

The primary outcome was psychotic distress assessed with the sum distress score of the CAARMS positive symptom subscales (range 0-100)^2^. Secondary outcomes included global and social functioning assessed with the SOFAS^29^ and SFS^30^, and symptom severity assessed with the BPRS^31^ and BNSS^32^ (see eMethods 2 for reliability measures and additional information on the measures). Other secondary outcomes were measured with the ESM, a structured, time-sampling diary technique^33,34^, and included momentary psychotic distress, momentary intensity of psychotic experiences, and momentary positive affect and negative affect (see eTable 1). Any serious adverse events were recorded throughout the entire study period.

**Table 1:**
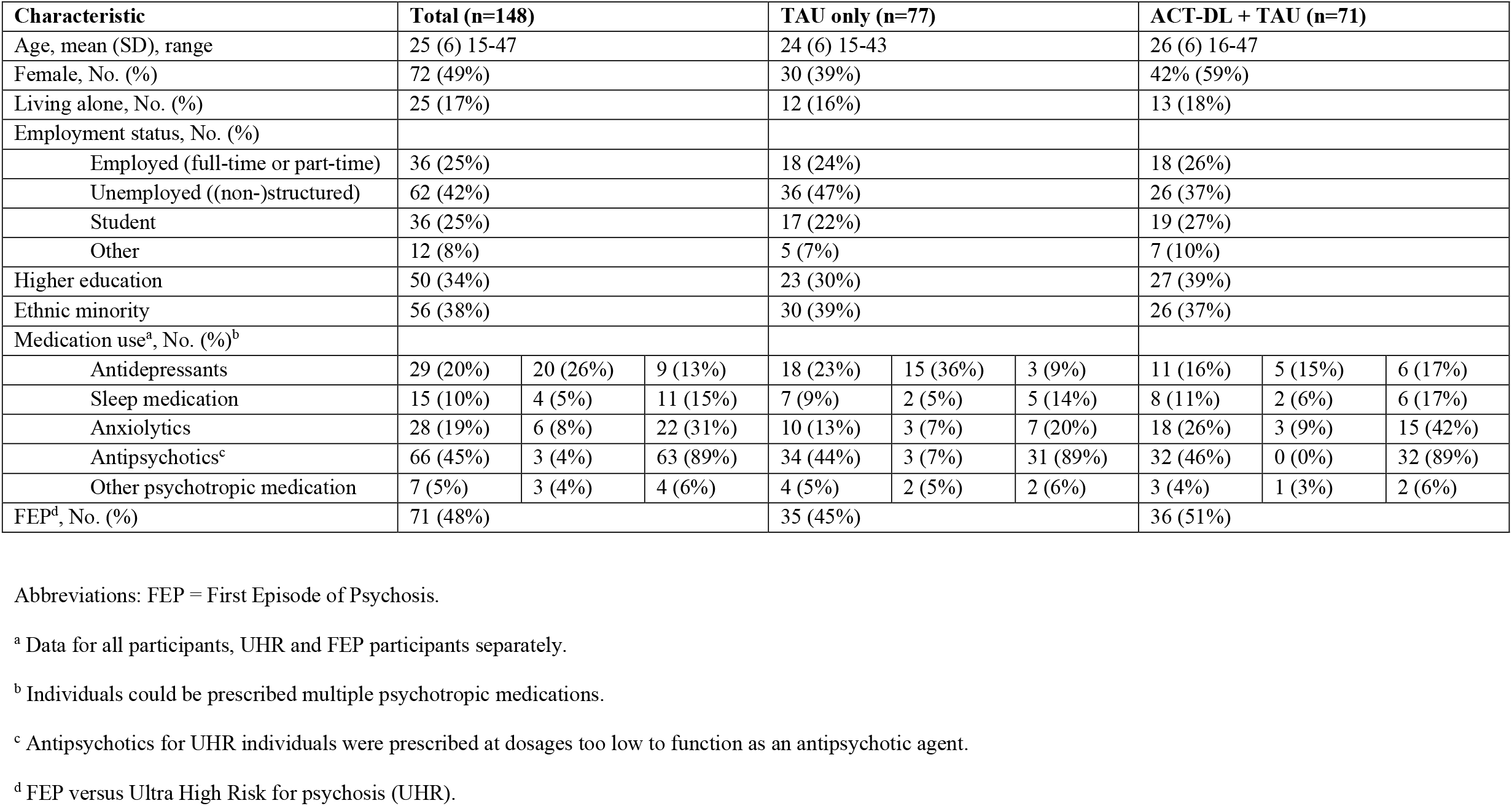
Baseline demographic and clinical characteristics of the intention-to-treat population.

| Characteristic | Total (n=148) |  |  | TAU only (n=77) |  |  | ACT-DL + TAU (n=71) |  |  |
| --- | --- | --- | --- | --- | --- | --- | --- | --- | --- |
| Age, mean (SD), range | 25 (6) 15-47 |  |  | 24 (6) 15-43 |  |  | 26 (6) 16-47 |  |  |
| Female, No. (%) | 72 (49%) |  |  | 30 (39%) |  |  | 42% (59%) |  |  |
| Living alone, No. (%) | 25 (17%) |  |  | 12 (16%) |  |  | 13 (18%) |  |  |
| Employment status, No. (%) |  |  |  |  |  |  |  |  |  |
| Employed (full-time or part-time) | 36 (25%) |  |  | 18 (24%) |  |  | 18 (26%) |  |  |
| Unemployed ((non-)structured) | 62 (42%) |  |  | 36 (47%) |  |  | 26 (37%) |  |  |
| Student | 36 (25%) |  |  | 17 (22%) |  |  | 19 (27%) |  |  |
| Other | 12 (8%) |  |  | 5 (7%) |  |  | 7 (10%) |  |  |
| Higher education | 50 (34%) |  |  | 23 (30%) |  |  | 27 (39%) |  |  |
| Ethnic minority | 56 (38%) |  |  | 30 (39%) |  |  | 26 (37%) |  |  |
| Medication use<sup>a</sup>, No. (%)<sup>b</sup> |  |  |  |  |  |  |  |  |  |
| Antidepressants | 29 (20%) | 20 (26%) | 9 (13%) | 18 (23%) | 15 (36%) | 3 (9%) | 11 (16%) | 5 (15%) | 6 (17%) |
| Sleep medication | 15 (10%) | 4 (5%) | 11 (15%) | 7 (9%) | 2 (5%) | 5 (14%) | 8 (11%) | 2 (6%) | 6 (17%) |
| Anxiolytics | 28 (19%) | 6 (8%) | 22 (31%) | 10 (13%) | 3 (7%) | 7 (20%) | 18 (26%) | 3 (9%) | 15 (42%) |
| Antipsychotics<sup>c</sup> | 66 (45%) | 3 (4%) | 63 (89%) | 34 (44%) | 3 (7%) | 31 (89%) | 32 (46%) | 0 (0%) | 32 (89%) |
| Other psychotropic medication | 7 (5%) | 3 (4%) | 4 (6%) | 4 (5%) | 2 (5%) | 2 (6%) | 3 (4%) | 1 (3%) | 2 (6%) |
| FEP<sup>d</sup>, No. (%) | 71 (48%) |  |  | 35 (45%) |  |  | 36 (51%) |  |  |

Abbreviations: FEP = First Episode of Psychosis.
<sup>a</sup> Data for all participants, UHR and FEP participants separately.
<sup>b</sup> Individuals could be prescribed multiple psychotropic medications.
<sup>c</sup> Antipsychotics for UHR individuals were prescribed at dosages too low to function as an antipsychotic agent.
<sup>d</sup> FEP versus Ultra High Risk for psychosis (UHR).

### Statistical analysis

Power simulation in R indicated that a sample size of 150 participants (75 per arm) would have 92% power to detect a medium effect size of d=0.5 at (at least) one of the post-intervention and follow-up time points when testing our primary hypothesis at alpha=0.05, while allowing for an expected attrition rate of 31% to follow-up.

Statistical analyses were conducted using Stata (version 14.2) and data were analyzed according to intention-to-treat principles. Multivariate multi-level regression models were fitted for primary and secondary outcomes separately with scores at post-intervention, 6-month and 12-month follow-up as the dependent variable. The independent variables included condition, time, group status (UHR or FEP), region, baseline score (grand-mean centered), a baseline score × time interaction, and a condition × time interaction. An omnibus test of no difference between the two conditions was performed at all three time points (Wald-type test with df=3 and alpha=0.05). Only if statistically significant, the three time-specific contrasts were examined (each tested at alpha=0.05), precluding the need for adjusting for multiple testing at the level of time-specific contrasts.

In the models with ESM outcomes, momentary psychotic experiences, positive and negative affect were included as dependent variables with the same independent variables as previously described. In the analysis of momentary psychotic distress (defined as the association between momentary psychotic experience and negative affect), negative affect was the dependent variable and momentary psychotic experiences (person-mean centered), a momentary psychotic experiences × time interaction, a momentary psychotic experiences × condition interaction, and a momentary psychotic experiences × time × condition interaction were added as additional independent variables to the model. For these models, an additional level of nesting was added with multiple ESM observations (level 1) being nested within time points (post-intervention, 6-month follow-up) (level 2) nested within subjects (level 3). We added level-3 random intercepts and slopes for the time points and set the variance-covariance matrix of these effects to unstructured. All models were fitted using restricted maximum likelihood (REML) estimation, allowing for the use of all available data under the assumption that data is missing at random and that all variables associated with missing values (see eTable 2) are included in the model^35,36^. For the subgroup analyses, see eMethods 3.

**Table 2:**
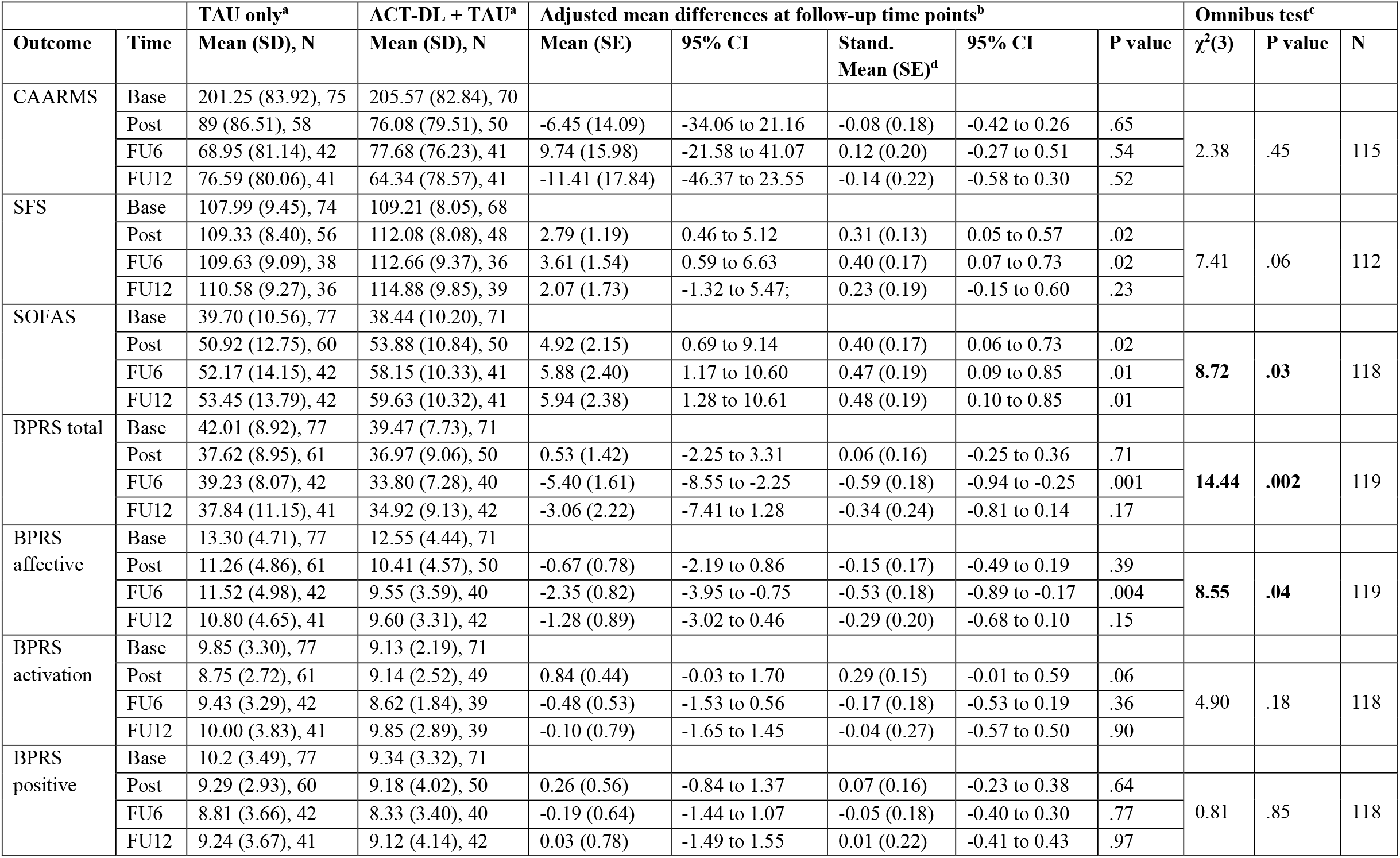
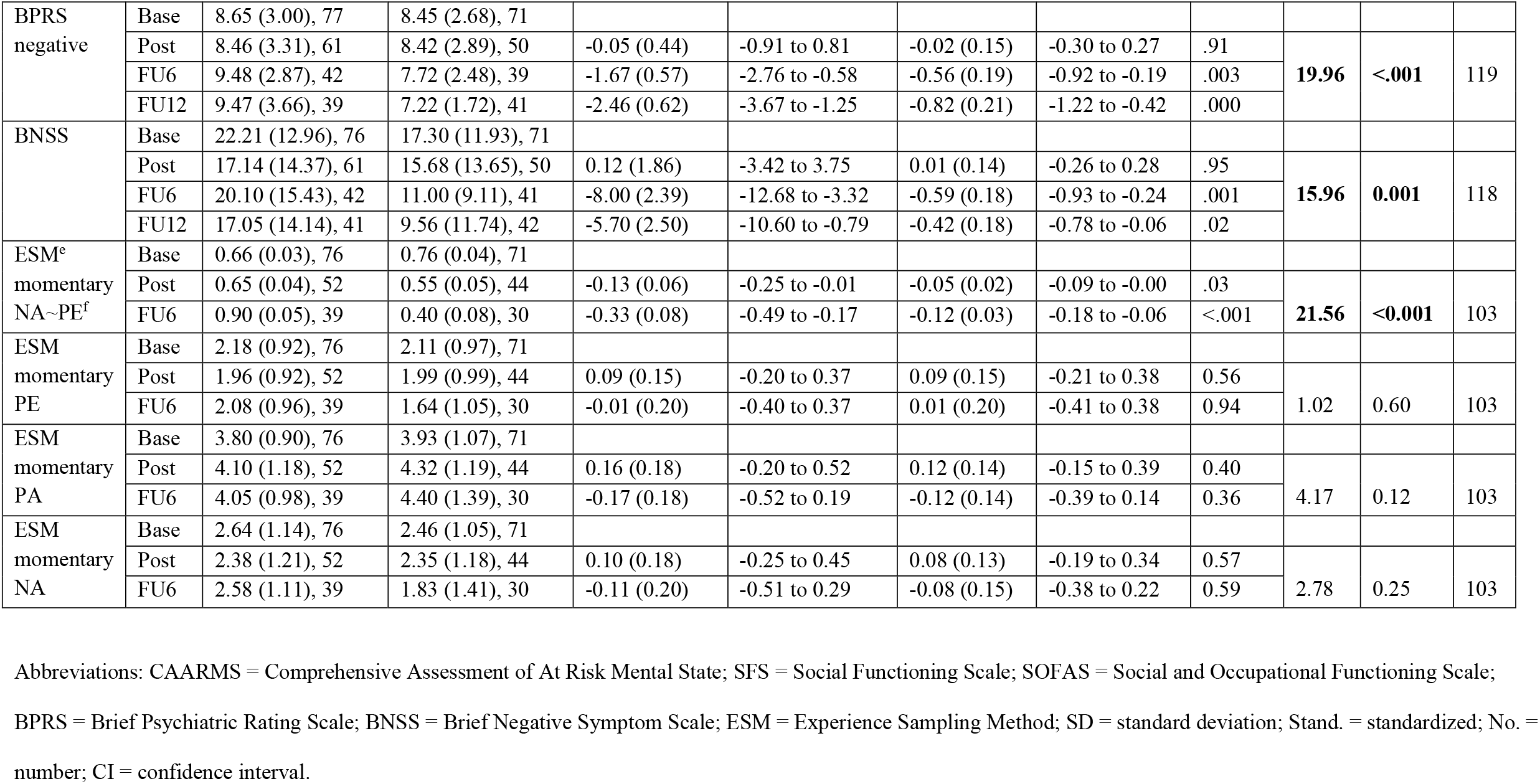
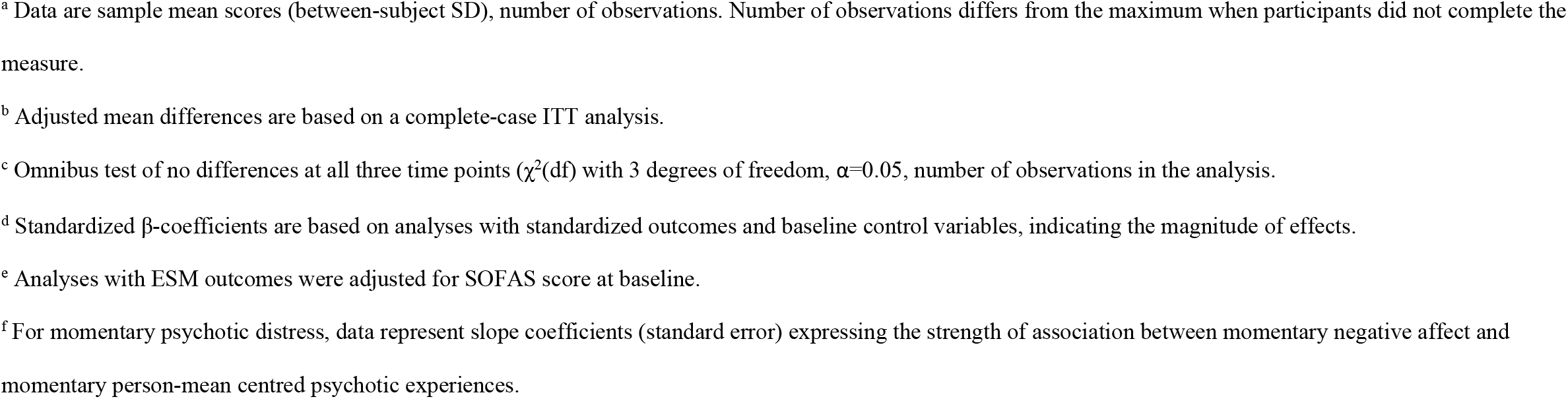
Primary and secondary outcomes at baseline, post-intervention, 6-month and 12-month follow-up.

|  |  | TAU only<sup>a</sup> | ACT-DL + TAU<sup>a</sup> | Adjusted mean differences at follow-up time points<sup>b</sup> |  |  |  |  | Omnibus test<sup>c</sup> |  |  |
| --- | --- | --- | --- | --- | --- | --- | --- | --- | --- | --- | --- |
| Outcome | Time | Mean (SD), N | Mean (SD), N | Mean (SE) | 95% CI | Stand. Mean (SE)<sup>d</sup> | 95% CI | P value | $\chi^2(3)$ | P value | N |
| CAARMS | Base | 201.25 (83.92), 75 | 205.57 (82.84), 70 |  |  |  |  |  |  |  |  |
|  | Post | 89 (86.51), 58 | 76.08 (79.51), 50 | -6.45 (14.09) | -34.06 to 21.16 | -0.08 (0.18) | -0.42 to 0.26 | .65 | 2.38 | .45 | 115 |
|  | FU6 | 68.95 (81.14), 42 | 77.68 (76.23), 41 | 9.74 (15.98) | -21.58 to 41.07 | 0.12 (0.20) | -0.27 to 0.51 | .54 |  |  |  |
|  | FU12 | 76.59 (80.06), 41 | 64.34 (78.57), 41 | -11.41 (17.84) | -46.37 to 23.55 | -0.14 (0.22) | -0.58 to 0.30 | .52 |  |  |  |
| SFS | Base | 107.99 (9.45), 74 | 109.21 (8.05), 68 |  |  |  |  |  |  |  |  |
|  | Post | 109.33 (8.40), 56 | 112.08 (8.08), 48 | 2.79 (1.19) | 0.46 to 5.12 | 0.31 (0.13) | 0.05 to 0.57 | .02 | 7.41 | .06 | 112 |
|  | FU6 | 109.63 (9.09), 38 | 112.66 (9.37), 36 | 3.61 (1.54) | 0.59 to 6.63 | 0.40 (0.17) | 0.07 to 0.73 | .02 |  |  |  |
|  | FU12 | 110.58 (9.27), 36 | 114.88 (9.85), 39 | 2.07 (1.73) | -1.32 to 5.47; | 0.23 (0.19) | -0.15 to 0.60 | .23 |  |  |  |
| SOFAS | Base | 39.70 (10.56), 77 | 38.44 (10.20), 71 |  |  |  |  |  |  |  |  |
|  | Post | 50.92 (12.75), 60 | 53.88 (10.84), 50 | 4.92 (2.15) | 0.69 to 9.14 | 0.40 (0.17) | 0.06 to 0.73 | .02 | 8.72 | .03 | 118 |
|  | FU6 | 52.17 (14.15), 42 | 58.15 (10.33), 41 | 5.88 (2.40) | 1.17 to 10.60 | 0.47 (0.19) | 0.09 to 0.85 | .01 |  |  |  |
|  | FU12 | 53.45 (13.79), 42 | 59.63 (10.32), 41 | 5.94 (2.38) | 1.28 to 10.61 | 0.48 (0.19) | 0.10 to 0.85 | .01 |  |  |  |
| BPRS total | Base | 42.01 (8.92), 77 | 39.47 (7.73), 71 |  |  |  |  |  |  |  |  |
|  | Post | 37.62 (8.95), 61 | 36.97 (9.06), 50 | 0.53 (1.42) | -2.25 to 3.31 | 0.06 (0.16) | -0.25 to 0.36 | .71 | 14.44 | .002 | 119 |
|  | FU6 | 39.23 (8.07), 42 | 33.80 (7.28), 40 | -5.40 (1.61) | -8.55 to -2.25 | -0.59 (0.18) | -0.94 to -0.25 | .001 |  |  |  |
|  | FU12 | 37.84 (11.15), 41 | 34.92 (9.13), 42 | -3.06 (2.22) | -7.41 to 1.28 | -0.34 (0.24) | -0.81 to 0.14 | .17 |  |  |  |
| BPRS affective | Base | 13.30 (4.71), 77 | 12.55 (4.44), 71 |  |  |  |  |  |  |  |  |
|  | Post | 11.26 (4.86), 61 | 10.41 (4.57), 50 | -0.67 (0.78) | -2.19 to 0.86 | -0.15 (0.17) | -0.49 to 0.19 | .39 | 8.55 | .04 | 119 |
|  | FU6 | 11.52 (4.98), 42 | 9.55 (3.59), 40 | -2.35 (0.82) | -3.95 to -0.75 | -0.53 (0.18) | -0.89 to -0.17 | .004 |  |  |  |
|  | FU12 | 10.80 (4.65), 41 | 9.60 (3.31), 42 | -1.28 (0.89) | -3.02 to 0.46 | -0.29 (0.20) | -0.68 to 0.10 | .15 |  |  |  |
| BPRS activation | Base | 9.85 (3.30), 77 | 9.13 (2.19), 71 |  |  |  |  |  |  |  |  |
|  | Post | 8.75 (2.72), 61 | 9.14 (2.52), 49 | 0.84 (0.44) | -0.03 to 1.70 | 0.29 (0.15) | -0.01 to 0.59 | .06 | 4.90 | .18 | 118 |
|  | FU6 | 9.43 (3.29), 42 | 8.62 (1.84), 39 | -0.48 (0.53) | -1.53 to 0.56 | -0.17 (0.18) | -0.53 to 0.19 | .36 |  |  |  |
|  | FU12 | 10.00 (3.83), 41 | 9.85 (2.89), 39 | -0.10 (0.79) | -1.65 to 1.45 | -0.04 (0.27) | -0.57 to 0.50 | .90 |  |  |  |
| BPRS positive | Base | 10.2 (3.49), 77 | 9.34 (3.32), 71 |  |  |  |  |  |  |  |  |
|  | Post | 9.29 (2.93), 60 | 9.18 (4.02), 50 | 0.26 (0.56) | -0.84 to 1.37 | 0.07 (0.16) | -0.23 to 0.38 | .64 | 0.81 | .85 | 118 |
|  | FU6 | 8.81 (3.66), 42 | 8.33 (3.40), 40 | -0.19 (0.64) | -1.44 to 1.07 | -0.05 (0.18) | -0.40 to 0.30 | .77 |  |  |  |
|  | FU12 | 9.24 (3.67), 41 | 9.12 (4.14), 42 | 0.03 (0.78) | -1.49 to 1.55 | 0.01 (0.22) | -0.41 to 0.43 | .97 |  |  |  |

Table 2 (continued): Primary and secondary outcomes at baseline, post-intervention, 6-month and 12-month follow-up.
| Outcome | Time | TAU only<sup>a</sup> | ACT-DL + TAU<sup>a</sup> | Adjusted mean differences at follow-up time points<sup>b</sup> |  |  |  |  | Omnibus test<sup>c</sup> |  |  |
| --- | --- | --- | --- | --- | --- | --- | --- | --- | --- | --- | --- |
| | | Mean (SD), N | Mean (SD), N | Mean (SE) | 95% CI | Stand. Mean (SE)<sup>d</sup> | 95% CI | P value | $\chi^2(3)$ | P value | N |
| BPRS negative | Base | 8.65 (3.00), 77 | 8.45 (2.68), 71 |  |  |  |  |  |  |  |  |
|  | Post | 8.46 (3.31), 61 | 8.42 (2.89), 50 | -0.05 (0.44) | -0.91 to 0.81 | -0.02 (0.15) | -0.30 to 0.27 | .91 | **19.96** | **<.001** | 119 |
|  | FU6 | 9.48 (2.87), 42 | 7.72 (2.48), 39 | -1.67 (0.57) | -2.76 to -0.58 | -0.56 (0.19) | -0.92 to -0.19 | .003 |  |  |  |
|  | FU12 | 9.47 (3.66), 39 | 7.22 (1.72), 41 | -2.46 (0.62) | -3.67 to -1.25 | -0.82 (0.21) | -1.22 to -0.42 | .000 |  |  |  |
| BNSS | Base | 22.21 (12.96), 76 | 17.30 (11.93), 71 |  |  |  |  |  |  |  |  |
|  | Post | 17.14 (14.37), 61 | 15.68 (13.65), 50 | 0.12 (1.86) | -3.42 to 3.75 | 0.01 (0.14) | -0.26 to 0.28 | .95 | **15.96** | **0.001** | 118 |
|  | FU6 | 20.10 (15.43), 42 | 11.00 (9.11), 41 | -8.00 (2.39) | -12.68 to -3.32 | -0.59 (0.18) | -0.93 to -0.24 | .001 |  |  |  |
|  | FU12 | 17.05 (14.14), 41 | 9.56 (11.74), 42 | -5.70 (2.50) | -10.60 to -0.79 | -0.42 (0.18) | -0.78 to -0.06 | .02 |  |  |  |
| ESM<sup>e</sup><br>momentary<br>NA~PE<sup>f</sup> | Base | 0.66 (0.03), 76 | 0.76 (0.04), 71 |  |  |  |  |  |  |  |  |
|  | Post | 0.65 (0.04), 52 | 0.55 (0.05), 44 | -0.13 (0.06) | -0.25 to -0.01 | -0.05 (0.02) | -0.09 to -0.00 | .03 | **21.56** | **<0.001** | 103 |
|  | FU6 | 0.90 (0.05), 39 | 0.40 (0.08), 30 | -0.33 (0.08) | -0.49 to -0.17 | -0.12 (0.03) | -0.18 to -0.06 | <.001 |  |  |  |
| ESM<br>momentary<br>PE | Base | 2.18 (0.92), 76 | 2.11 (0.97), 71 |  |  |  |  |  |  |  |  |
|  | Post | 1.96 (0.92), 52 | 1.99 (0.99), 44 | 0.09 (0.15) | -0.20 to 0.37 | 0.09 (0.15) | -0.21 to 0.38 | 0.56 | 1.02 | 0.60 | 103 |
|  | FU6 | 2.08 (0.96), 39 | 1.64 (1.05), 30 | -0.01 (0.20) | -0.40 to 0.37 | 0.01 (0.20) | -0.41 to 0.38 | 0.94 |  |  |  |
| ESM<br>momentary<br>PA | Base | 3.80 (0.90), 76 | 3.93 (1.07), 71 |  |  |  |  |  |  |  |  |
|  | Post | 4.10 (1.18), 52 | 4.32 (1.19), 44 | 0.16 (0.18) | -0.20 to 0.52 | 0.12 (0.14) | -0.15 to 0.39 | 0.40 | 4.17 | 0.12 | 103 |
|  | FU6 | 4.05 (0.98), 39 | 4.40 (1.39), 30 | -0.17 (0.18) | -0.52 to 0.19 | -0.12 (0.14) | -0.39 to 0.14 | 0.36 |  |  |  |
| ESM<br>momentary<br>NA | Base | 2.64 (1.14), 76 | 2.46 (1.05), 71 |  |  |  |  |  |  |  |  |
|  | Post | 2.38 (1.21), 52 | 2.35 (1.18), 44 | 0.10 (0.18) | -0.25 to 0.45 | 0.08 (0.13) | -0.19 to 0.34 | 0.57 | 2.78 | 0.25 | 103 |
|  | FU6 | 2.58 (1.11), 39 | 1.83 (1.41), 30 | -0.11 (0.20) | -0.51 to 0.29 | -0.08 (0.15) | -0.38 to 0.22 | 0.59 |  |  |  |

Abbreviations: CAARMS = Comprehensive Assessment of At Risk Mental State; SFS = Social Functioning Scale; SOFAS = Social and Occupational Functioning Scale;
BPRS = Brief Psychiatric Rating Scale; BNSS = Brief Negative Symptom Scale; ESM = Experience Sampling Method; SD = standard deviation; Stand. = standardized; No. = number; CI = confidence interval.
<sup>a</sup> Data are sample mean scores (between-subject SD), number of observations. Number of observations differs from the maximum when participants did not complete the measure.
<sup>b</sup> Adjusted mean differences are based on a complete-case ITT analysis.
<sup>c</sup> Omnibus test of no differences at all three time points ( $\chi^2(df)$ with 3 degrees of freedom, $\alpha=0.05$ , number of observations in the analysis).
<sup>d</sup> Standardized β-coefficients are based on analyses with standardized outcomes and baseline control variables, indicating the magnitude of effects.
<sup>e</sup> Analyses with ESM outcomes were adjusted for SOFAS score at baseline.
<sup>f</sup> For momentary psychotic distress, data represent slope coefficients (standard error) expressing the strength of association between momentary negative affect and momentary person-mean centred psychotic experiences.

## Results

Of the 196 individuals assessed for eligibility (fig 1), 148 participants were randomized to ACT-DL+TAU (n=71) or to TAU (n=77 with n=27 receiving CBTp+TAU). Attrition rates for the primary outcome at post-intervention assessment were 21 (30%) participants in ACT-DL+TAU and 17 (22%) in TAU. Attrition rate was higher than expected at 6-month and 12-month follow-up (see Figure 1).

**Figure 1.**
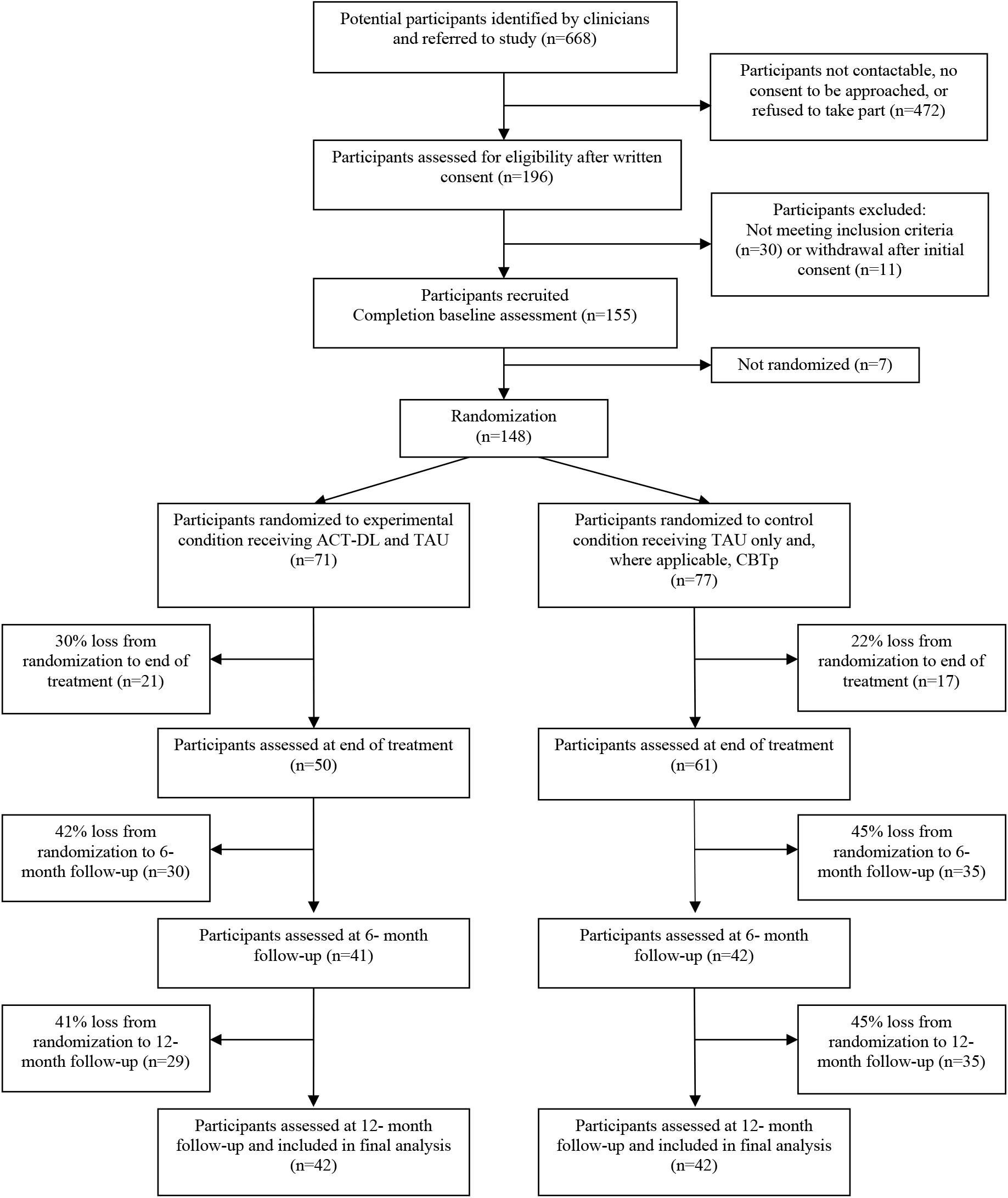
Study flowchart Abbrevations: ACT-DL, Acceptance and Commitment Therapy in Daily Life; TAU, treatment as usual; CBTp, Cognitive Behavioural Therapy for psychosis.

The sample included 77 individuals with UHR and 71 with FEP (see Table 1). The sample was nearly equally divided between men and women, with a slightly larger proportion of women in ACT-DL+TAU compared to TAU. The majority of participants lived with others, were employed or student, and did not obtain higher education. Almost 40% of participants in each condition were from an ethnic minority group.

The mean fidelity score was 10.7 (SD=1.5; range [6.5-12.5), demonstrating excellent adherence to the ACT-DL manual. Patients participated on average in 6 (SD 3) out of 8 sessions. Unblinding occurred in 21 (14%) of the participants, 16 in ACT-DL+TAU and 5 in TAU.

Table 2 shows descriptive statistics, adjusted mean differences and standardized β-coefficients indicating magnitude of effects for primary and secondary outcomes. There was no evidence of a greater reduction in CAARMS distress in ACT-DL+TAU compared to TAU. However, ACT-DL+TAU showed greater improvement in global functioning (SOFAS) at post-intervention, 6-month, and 12-month follow-up, but failed to reach statistical significance in social functioning (SFS). Moreover, greater reductions in BPRS symptom scores were found in ACT-DL+TAU for the total and affective symptom scale at 6-month follow-up as well as for the negative symptom scale at 6-month and 12-month follow-up (figure 2). In addition, compared with TAU, ACT-DL+TAU was associated with greater reductions in BNNS negative symptoms at 6-month and 12-month follow-up. There were no significant effects on momentary positive affect, negative affect and psychotic experiences. However, there was strong evidence of greater reductions in momentary distress associated with psychotic experiences in ACT-DL+TAU compared to TAU. Specifically, this showed that the reduction in the *association* between momentary psychotic experiences and momentary negative affect was greater in ACT-DL+TAU than TAU at both post-intervention and 6-month follow-up.

**Figure 2.**
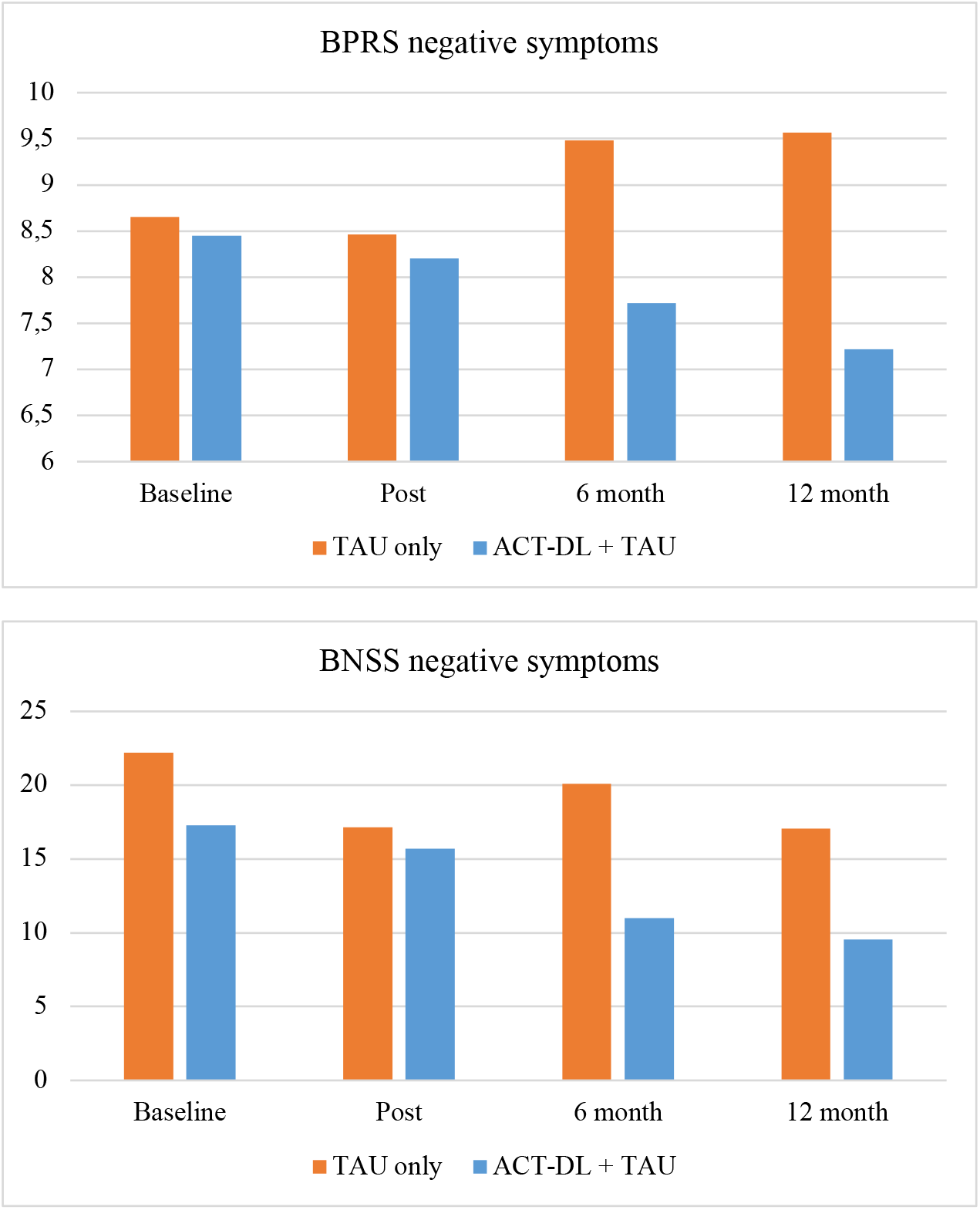
Differences in treatment effects on BPRS negative symptoms (top) and BNSS negative symptoms (bottom) at post-intervention, 6-month and 12-month follow-up.

The planned subgroup analysis comparing the between-condition effect in UHR versus FEP individuals yielded no significant results (eTable 3 and Figure 3). Likewise, our exploratory analysis comparing ACT-DL+TAU versus TAU with ACT-DL+TAU versus CBTp+TAU did not reach significance (eTable 4). Finally, serious adverse events did not occur as a result of the therapy.

**Figure 3.**
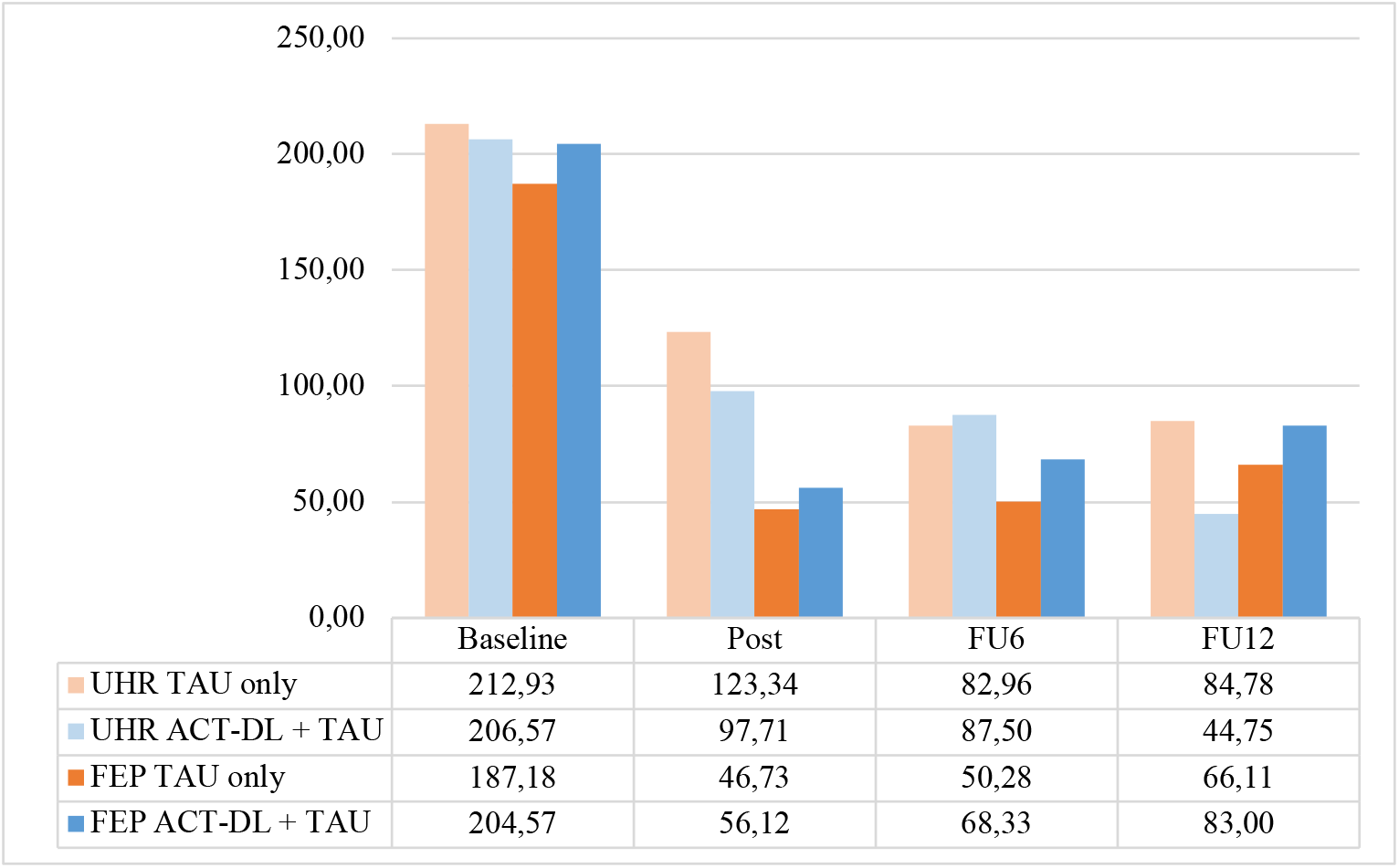
Differences in treatment effects on CAARMS distress scores in UHR versus FEP individuals at post-intervention, 6-month and 12-month follow-up.

## Discussion

No effect was found of ACT-DL+TAU compared to TAU on the primary outcome measure of psychotic distress in individuals at the early stages of psychosis. However, substantial improvements in the experimental condition were found for the secondary outcome measures, that is, for momentary psychotic distress, negative symptoms at 6-and 12-month follow-up, evident in two separate measures, as well as for global functioning at post-intervention, 6-and 12-month follow-up.

The lack of significant findings on psychotic distress seems mainly due to the fact that TAU was equally successful as ACT-DL+TAU in reducing psychotic distress over time. This is in line with a recent study showing that distress in UHR individuals tended to sharply decline in the first three months after inclusion in an intervention trial^37^, irrespective of the intervention arm, indicating that distress in these early phases of illness may be ameliorated by just monitoring and having someone to talk to. This seems supported by the literature, as a recent comprehensive meta-analysis, indeed, found no differential effect of any psychological nor pharmacological intervention on psychotic distress in the early phases of psychosis^10^. However, ACT-DL+TAU is associated with larger improvements in self-reported psychotic distress in daily life compared to TAU, indicating that more fine-grained and self-reported real-life measures may capture more subtle changes in psychotic distress as it unfolds in daily life.

In contrast to the null-finding on the primary outcome, we found a consistently stronger effect of ACT-DL+TAU compared to TAU in the models of negative symptoms and global functioning, particularly after 6 and 12 months. This is an important finding as both negative symptoms and social functioning are considered the hardest to treat. Although some recent meta-analytic evidence showed small effects of cognitive behaviorally informed interventions on negative symptoms^38,39^ and functioning^38^ in individuals with psychosis, two meta-analyses specifically focusing on youth at clinical high risk for psychosis concluded that, currently, no effective treatments are available for either improving functioning^40^ or ameliorating negative symptoms^41^ in the early stages of psychosis. Yet, negative symptoms are considered the most disabling symptoms^42,43^, being closely associated with lower levels of functioning and higher risk for a more disabling course of illness. Improving both negative symptoms and functioning in these early phases of psychosis is thus very promising for prevention of further deterioration towards severe and recurrent symptoms in later stages of illness.

The finding that ACT-DL+TAU is particularly relevant for improving negative symptoms and global functioning is unexpected, given the current state-of-the-art of ACT therapy in psychosis. One meta-analysis found no effect of acceptance-based interventions on either negative symptoms or functioning^9^ and another study did not find a significant difference between second-(CBT) and third-(ACT) wave interventions on negative symptoms or functioning^38^. However, there are a number of reasons that may explain this discrepancy. First, this is the largest trial examining ACT for psychosis. Previous studies may have been underpowered to detect significant effects. Second, this study used a 12-month follow-up to examine both the immediate and longer-term effects. In contrast, most previous ACT studies used no or a short-term follow-up^15^. Third, this is the first RCT focusing on the early stages of psychosis. Acceptance of distress in these early stages may have contributed to patients being less experientially avoidant in dealing with overwhelming symptoms, which may explain the effects on negative symptoms, while ‘committed action’ may have been particularly instrumental in improving functioning. Fourth, the current study used a blended care intervention, combining face-to-face ACT therapy with the ACT-DL EMI. Although the current study did not evaluate added benefit of ACT-DL over ACT only, the findings of continuous improvement of both functioning and negative symptoms over time may indicate the added value of the EMI component. Indeed, whereas long-term effects of psychological interventions tend to either stay the same^44^ or decline over time^45^, a previous EMI in individuals with depression showed a similar pattern of continuous increase in effect up to 24 weeks post-intervention^46^. This might be explained by the fact that EMIs are particularly tuned to applying skills in everyday situations and, thus, towards developing new habits that will increasingly have an effect over time^17^. Further research is needed to demonstrate and disentangle the effects of ACT versus ACT-DL in this population, as well as to identify the contributing components to the long-term effectiveness on negative symptoms and functioning.

The strengths of the current study are the sample size, the use of an active control condition in a subset of participants (including both TAU and structured CBTp), the rigorous trial approach and open science practices (with the study protocol published and the detailed postregistration of the analysis plan), the use of a blended care intervention and the wide scope of primary and secondary outcomes, measured, in part, using ESM. The results of this study also need to be interpreted in light of a number of limitations. First, the control condition consists of both non-manualized TAU and TAU including manualized CBTp. Therefore, we did not control for the effect of receiving structured psychotherapy *per se* in all participants allocated to TAU. However, TAU in the different centers usually included some form of psychotherapy. Second, the sample included both FEP and UHR individuals. Although both groups represent the early stages of psychosis and are considered temporally and phenomenologically continuous, they do present different stages of illness according to the staging model of psychosis. Indeed, most early intervention studies focus particularly on the UHR group. We randomized the groups stratified by UHR versus FEP, making sure that both groups were equally distributed over treatment conditions. Furthermore, the planned subgroup analysis showed no differences between UHR and FEP. Third, whereas use of antipsychotic medication was an exclusion criterion for UHR, most of the FEP were prescribed antipsychotics. We documented this in the sample characteristics. Furthermore, over the course of treatment (including the 6-and 12-month follow-up) individuals could naturally be prescribed antipsychotic medication if clinically indicated, but given the randomization this factor was, as other potential confounders, balanced across conditions. Fourth, ACT-DL included both a face-to-face and an EMI intervention. However, based on this trial, we cannot disentangle the working components.

To conclude, the blended care approach of face-to-face ACT with the ACT-DL EMI provides a promising avenue for the treatment of momentary psychotic distress and negative symptoms while also improving functioning, in individuals with UHR and FEP. This is an important finding as effective treatment at the early stages of both of these problems, may prevent deterioration to more severe stages of illness.

## Supporting information

CONSORT checklist

## Data Availability

Deidentified data are available upon request through a data access system, Data cuRation for OPen Science (DROPS), administered via REDCap at the Center for Contextual Psychiatry, KU Leuven. Interested researchers can submit an abstract, which is subject to review by the research team to ensure there is no overlap with existing projects. Following abstract approval, a variable access request is submitted and researchers are required to postregister their analysis plan. A dataset containing only variables required for the proposed analysis is then released to the researchers by a data manager, along with a time- and date-stamped receipt of data access.

https://osf.io/5qfwe/?view_only=4062ccf54b0d4161a0e481aa80e76e78

## Contributors

**Inez Myin-Germeys**: Conceptualization, Methodology, Writing – Original Draft (introduction and discussion), Writing – Review & Editing, Visualization, Supervision, Project administration, Funding acquisition. **Evelyne van Aubel**: Conceptualization, Methodology, Software, Formal analysis, Investigation (recruitment and assessment), Data curation, Writing – Original draft (method section and results), Writing – Review & Editing, Visualization. **Ginette Lafit**: Software, Formal analysis, Writing – Review & Editing. **Wolfgang Viechtbauer**: Methodology, Software, Writing – Review & Editing. **Thomas Vaessen**: Conceptualization, Methodology, Investigation (recruitment and assessment), Writing – Review & Editing. **Henrietta Steinhart**: Conceptualization, Methodology, Investigation (recruitment and assessment), Writing – Review & Editing. **Annelie Klippel**: Conceptualization, Methodology, Investigation (recruitment and assessment), Writing – Review & Editing. **Tim Batink**: Resources (supervision to trial therapists). **Ruud van Winkel**: Investigation (recruitment), Writing – Review & Editing. **Mark van der Gaag**: Conceptualization, Investigation (recruitment), Writing – Review & Editing. **Therese van Amelsvoort**: Investigation (recruitment), Writing – Review & Editing. **Machteld Marcelis**: Investigation (recruitment), Writing – Review & Editing. **Frederike Schirmbeck**: Conceptualization, Methodology, Investigation (recruitment), Writing – Review & Editing. **Lieuwe De Haan:** Conceptualization, Methodology, Writing – Review & Editing. **Ulrich Reininghaus**: Conceptualization, Methodology, Writing – Original draft (method section and results), Writing – Review & Editing, Supervision, Project administration (PI of the NWO VENI grant), Funding acquisition.

## Data sharing

Deidentified data are available upon request through a data access system, ‘Data cuRation for OPen Science’ (DROPS), administered via REDCap at the Center for Contextual Psychiatry, KU Leuven. Interested researchers can submit an abstract, which is subject to review by the research team to ensure there is no overlap with existing projects. Following abstract approval, a variable access request is submitted and researchers are required to postregister their analysis plan. A dataset containing only variables required for the proposed analysis is then released to the researchers by a data manager, along with a time-and date-stamped receipt of data access.

## Acknowledgments

This work was supported by an ERC Consolidator Grant (ERC – 2012 – StG, project 309767 – INTERACT) to IMG as well as a NWO VENI Grant (no. 451 – 13 – 022) and DFG Heisenberg professorship (no. 389624707) to UR. We thank all the participating health services in Amsterdam (Academic Medical Centre, Arkin Basis GGZ), The Hague (Parnassia, PsyQ), Maastricht/Eindhoven (Mondriaan, Virenze, GGZE), Flemish-Brabant (UPC KU Leuven, VDIP Antwerp, Sint-Annendael, PCM Mortsel), and East/West Flanders (OLV Brugge, Karus Melle, VDIP Sint Niklaas). We thank all research coordinators (Silke Apers, Nele Volbragt, Wendy Beuken), research assistants (Dieuwke Siegmann, Davinia Verhoeven, Anna de Zwart, Iris de Wit, Lore Depraetere, Tessa Biesemans, Lotte Hendriks), and data managers (Martien Wampers, Jolien Bynens) past and present who were involved in the INTERACT trial. We also like to thank all individuals who participated in the study and were essential for its successful completion.

## Footnotes

1 The current study was postregistered, meaning that we registered our analysis plan after we had collected data, however before we had any access to the data. The main postregistration for the INTERACT study is available here https://osf.io/du2bn/?view_only=ec22ed02651441349e1bb1242cfc712c.The postregistration for the current study is embedded as a file within the main registration and can be accessed here https://osf.io/5qfwe/?view_only=4062ccf54b0d4161a0e481aa80e76e78.

