## Supplementary material for "Efficacy of Acceptance and Commitment Therapy in Daily Life (ACT-DL) in early psychosis: Results from the multi-center INTERACT randomized controlled trial"

#### Supplementary Methods

**eMethod 1. Transparent changes document.**

This study was post-registered at the OSF website (https://osf.io/du2bn/?view_only=f0d1e1d6b5fe498d8d143e8d80323090). We made **two minor deviations** from this report which we report here in detail as a "transparent changes" supplement and which we have made available on the OSF website as well.

The first deviation concerns the calculation of the ESM outcome "**momentary intensity of psychotic experiences**". In our original report, we stated that we would average the following ten ESM-questionnaire items to calculate this variable: "*I feel suspicious*", "*I feel unreal*", "*my thoughts cannot be expressed*", "*my thinking if confused*", "*I cannot get rid of my thoughts*", "*my thoughts are being influenced*", "*I hear voices*", "*I see appearances*", "*I hear things that aren't really there*", "*I'm afraid to lose control*". After the data had been accessed, we conducted an exploratory factor analysis on these items to assess construct validity. This analysis yielded two subscales (i.e. a disorganized thoughts scale (factor 1) and a hallucinations scale (factor 2)). The items "*my thoughts are being influenced*" and "*I'm afraid to lose control*" did not reach a sufficient loading of .40[1] on any of these factors, and were therefore excluded. We decided to perform all analyses with a total 8-item "momentary psychotic experiences" scale.

The second deviation concerns the data **exclusion rule for our ESM data**. As mentioned in the manuscript, there were three independent periods of six ESM assessment days throughout the study: at baseline, post-intervention, and 6-month follow-up. Participants were excluded from randomization at baseline if they did not fill out at least 25% (i.e. 15 out of 60 beeps) of the notifications during this 6-day period. It is of note that this rule was only instigated from December 2017 onwards. In our original post-registration report, we stated that "*data of participants who [had] less than 25% of the beeps at baseline [would] be excluded from the analyses*", and that "*participants need[ed] a minimum of 25% of the beeps at either post-intervention or 6-month follow-up to be included as well*". However, upon accessing the ESM dataset, we encountered unforeseen issues with the data. For one thing, we found that those individuals who had been randomized before December 2017 often had fewer than 25% of the notifications at baseline within a window of six days. For another, some individuals had used the PsyMate™ more than the intended six days within some assessment periods to reach the arbitrary cut-off of 25% of the notifications. As such, following our post-registered data exclusion rule would lead to a large amount of important ESM data lost, which we wanted to avoid at all cost. At the same time, it was very important to us to follow the intended six-day protocol, making each ESM assessment period of each participant comparable to one another. As such, we adjusted our data exclusion approach accordingly. That is, for each participant, at each time point, we calculated the window of six days in which this participant filled in the highest number of beeps and subsequently excluded all data that fell outside this window. Second, we checked with a multilevel logistic regression model whether baseline characteristics and condition predicted missingness at the level of the ESM questionnaire notifications and if so, we controlled for these predictors in all subsequent ESM analyses (see eTable 2 for more information).

**eMethod 2. Measures and procedures.**

- Choice of primary outcome measure

The primary outcome was distress associated with psychotic experiences measured with the sum distress score of the CAARMS positive symptom subscales (range 0-100)>. The CAARMS is a semi-structured interview gathering detailed information on the intensity, frequency, and emotional distress of various positive symptoms to detect whether individuals meet UHR or FEP criteria, with good psychometric properties[2]. It has been used to assess psychotherapy-induced changes in psychotic distress in both UHR and FEP individuals[3], thus providing a high-quality instrument that can be used across the different stages of early psychosis. The instrument has been widely adopted and is freely available in different languages. Thorough clinical training before using the instrument is necessary.

- ESM measures

There were three independent periods of six ESM days throughout the study: at baseline, post-intervention, and at 6-month follow-up. At baseline, participants were excluded from randomization if they did not fill out at least 25% (i.e. 15 out of 60 beeps) of the notifications during a 6-day period from December 2017 onwards. If participants did not fill out enough beeps after the first 6 days, they were given an additional 6 days of ESM assessment. Within each ESM assessment period, the PsyMate™ prompted participants with an ESM questionnaire 10 times a day at random moments within set blocks of time. After participants received a notification, the questionnaire remained available for 15 minutes, after which it disappeared. Participants were provided with a 30-minute extensive ESM briefing. In this briefing, they were instructed to carry the smartphone with them at all times, to answer as many notifications as possible, and to fill out the questionnaire while thinking about the moment right before the notification went off (meaning that the activity-related items had to be answered independently from the actual ESM itself). Once the briefing was completed, participants went through a demo questionnaire together with one of the assessors to make sure that every item was comprehensible. For more information on the ESM measures, please refer to eTable 1.

- Inter-rater reliability meetings

We conducted regular reliability meetings to assess interrater reliability for audiotaped scores on all clinical interviews, including the CAARMS (intensity and frequency score), SOFAS (total score), BPRS (total and subscale scores), and BNSS (total score). Intra-class correlation coefficients were calculated to examine interrater reliability using the iccNA command in R. Interrater reliability analyses yielded the following scores: 0.79 to 0.96 for CAARMS intensity and 0.81 to 0.93 for frequency scores, 0.67 for the SOFAS, 0.84 to 0.95 for BPRS (sub)scales, and 0.87 for the BNSS, showing sufficient agreement in all scales.

- Settings were data were collected

Data were collected at clinical sites of five regions: (1) Amsterdam (Academic Medical Centre, Arkin Basis GGZ), (2) The Hague (Parnassia/PsyQ), (3) Maastricht/Eindhoven (Mondriaan, Virenze, GGZE) (all in the Netherlands), (4) Flemish-Brabant (Leuven (UPC KU Leuven), Antwerp (VDIP), Diest (Sint-Annendael), Mortsel (PCM)), and (5) East/West Flanders (Brugge (OLV), Melle (Karus), Sint Niklaas (VDIP)) (all in Belgium).

- Treatment fidelity

Treatment fidelity was rated based on a random selection of audiotapes of three training sessions per participant using an adherence checklist covering all core ACT and ACT-DL app components within each session>. Ratings were based on the extent to which the component was addressed in each session (0 no, 1 to some extent, 2 yes), with 7 components to score in the sessions on contact with the present moment and values, and 6 components in all other sessions. We calculated a mean fidelity score (range 0 – 12.6).

**eMethod 3. Statistical analysis**

Statistical analyses were specified in the published protocol> and post-registered analysis plan.

- Within-subject clustering of repeated measures.

Within-subject clustering of repeated measures was taken into account by allowing residuals within subjects to be correlated with a completely unstructured variance-covariance matrix. For the models with ESM outcomes, an additional level of nesting was added with multiple ESM observations (level 1) being nested within time points (post-intervention, 6-month follow-up) (level 2) and time points as nested within subjects (level 3). We added level-3 random intercepts and slopes for the time points and set the variance-covariance matrix of these effects to unstructured. Within each level-2 time point (post-intervention and 6-month follow-up), level-1 within-subject residual errors were modelled to have an autoregressive (AR) structure (of the exponential type), which allows these models to account for unequally spaced time values.

- Estimation method and missing data.

All models were fitted using restricted maximum likelihood (REML) estimation, allowing for the use of all available data under the assumption that data is missing at random and that all variables associated with missing values are included in the model. To test for the latter, we fitted multilevel logistic regression models to examine if baseline characteristics were associated with missingness in the primary outcome or with missingness in secondary ESM outcomes at follow-up. If significant, these characteristics were added to the models as covariates. We found no association between baseline variables, condition, and missingness of the primary outcome measure. However, as global functioning (SOFAS) was significantly associated with missingness in secondary ESM outcome measures, analyses on these outcomes were controlled for this variable (eTable 2).

- Subgroup analysis: UHR versus FEP.

To test whether the effect of condition on CAARMS distress score differed between UHR and FEP participants, we added time × group, condition × group, and condition × time × group interactions to the model (planned subgroup analysis). Again, an omnibus test of no group difference (UHR vs. FEP) in the condition effect (which is the difference between ACT+TAU vs. TAU) at all three time points was performed and, only if statistically significant, time-specific contrasts were examined.

- Exploratory analysis: ACT-DL+TAU versus TAU only versus CBTp.

In a more exploratory sensitivity analysis, we probed our findings further to investigate whether the effect of ACT-DL+TAU versus TAU (difference 1) on CAARMS distress score was different from the effect of ACT-DL+TAU versus CBTp (difference 2) by including a 3-level factor variable for condition in the model. Also, in this model, we performed an omnibus test of no difference between ACTDL+ TAU and TAU versus ACT-DL+TAU and CBTp at all three time points before looking into time-specific contrasts.

#### Supplementary tables

**eTable 1. ESM measures.**

### ESM measures.

| Domain | ESM measure |
|---|---|
| Distress associated with psychotic experiences | Distress associated with psychotic experiences was operationalized as the within-subject association (i.e. the coefficient) between momentary negative affect and momentary intensity of psychotic experiences (person-mean centred) at the notification level, with the former being an outcome variable and the latter being a predictor variable. |
| Psychotic experiences | In line with previous ESM studies5–7, an ESM psychosis measure was used to assess intensity of psychotic experiences (PE). This measure consisted of 8 items: “I feel suspicious”, “I feel unreal”, “my thoughts cannot be expressed”, “my thinking is confused”, “I cannot get rid of my thoughts”, “I hear voices”, “I see appearances”, “I hear things that aren’t really there”. All items were rated on a 7-point Likert scale ranging from 1 (not at all) to 7 (very much). |
| Negative affect and positive affect | Negative affect (NA) and positive affect (PA) were assessed with a 6-item and a 4-item measure respectively. As for the first, participants were asked to indicate to what extent they felt lonely, insecure, anxious, irritated, down, and guilty on a 7-point Likert scale ranging from 1 (not at all) to 7 (very much). As for the latter, participants indicated how cheerful, relaxed, content, and enthusiastic they felt on that same scale. |

**eTable 2. Relation between missingness and randomised group and baseline scores.**

#### Relation between missingness and randomised group and baseline scores.

| | Missingness primary outcome[a] | | | Missingness ESM outcomes[b] | | |
|---|---|---|---|---|---|---|
| | **Coeff. (SE)** | **95% CI** | **P value** | **Coeff. (SE)** | **95% CI** | **P value** |
| Condition | -0.04 (0.34) | -0.71 to 0.63 | 0.90 | -0.23 (0.16) | -0.54 to 0.09 | 0.16 |
| CAARMS | -0.00 (0.00) | -0.01 to 0.00 | 0.07 | 0.00 (0.00) | -0.00 to 0.00 | 0.14 |
| SFS | 0.00 (0.02) | -0.05 to 0.05 | 0.97 | 0.01 (0.01) | -0.01 to 0.03 | 0.42 |
| SOFAS | -0.00 (0.02) | -0.04 to 0.03 | 0.87 | -0.02 (0.01) | -0.04 to -0.00 | 0.013 |
| BPRS | 0.03 (0.03) | -0.02 to 0.08 | 0.30 | -0.02 (0.01) | -0.04 to 0.00 | 0.09 |
| BNSS | -0.01 (0.02) | -0.04 to 0.03 | 0.71 | -0.01 (0.01) | -0.02 to 0.01 | 0.34 |
| NA | -0.12 (0.28) | -0.68 to 0.43 | 0.66 | 0.17 (0.13) | -0.10 to 0.43 | 0.22 |
| PA | 0.09 (0.20) | -0.30 to 0.49 | 0.64 | -0.00 (0.10) | -0.20 to 0.19 | 0.98 |
| PE | 0.06 (0.31) | -0.55 to 0.67 | 0.85 | 0.15 (0.16) | -0.16 to 0.45 | 0.34 |

Abbreviations: Coeff. = b-coefficient based on a multilevel logistic regression model; SE = standard error; 95% CI = 95% confidence interval of the b-coefficient; CAARMS = Comprehensive Assessment of At Risk Mental State; SFS = Social Functioning Scale; SOFAS = Social and Occupational Functioning Scale; BPRS = Brief Psychiatric Rating Scale; BNSS = Brief Negative Symptom Scale; ESM = Experience Sampling Method; NA = negative affect; PA = positive affect; PE = psychotic experiences.

[a] Missingness in the primary outcome (i.e. CAARMS distress score).

[b] Missingness in the ESM outcomes due to notification non-compliance.

**eTable 3. UHR and FEP subgroup analysis.**

#### Planned subgroup analysis of treatment effect on CAARMS distress scores in UHR and FEP individuals at post-intervention, 6-month, and 12-month follow-up.

| | | | | | Omnibus test | | |
|---|---|---|---|---|---|---|---|
| **Effects[a]** | **Time** | **Coefficient (SE)** | **95% CI** | **P value** | **$\chi^2(3)$ [b]** | **P value** | **No.** |
| Time × condition | Post | -21.95 (19.44) | -60.04 to 16.15 | 0.26 | | | |
| | FU6 | 8.32 (22.25) | -35.29 to 51.93 | 0.71 | | | |
| | FU12 | -36.91 (23.43) | -82.83 to 9.00 | 0.12 | | | |
| Time × group | Post | -58.67 (19.86) | -97.60 to -19.74 | 0.00 | | | |
| | FU6 | -15.96 (23.45) | -61.92 to 30.01 | 0.50 | | | |
| | FU12 | -1.36 (24.84) | -50.04 to 47.32 | 0.96 | | | |
| Condition × time × group | Post | 33.84 (27.80) | -20.64 to 88.33 | 0.22 | 4.50 | 0.21 | 115 |
| | FU6 | 1.02 (32.39) | -62.47 to 64.50 | 0.98 | | | |
| | FU12 | 47.79 (34.18) | -19.22 to 114.79 | 0.16 | | | |

Abbreviations: SE = standard error; CI = confidence interval; No. = number

a Effects represent the main effect of condition, group, and the condition × group interaction at various time points within the study.

b The omnibus test tests whether treatment effects are different in UHR versus FEP individuals across all three time points, reflected by the 3 degrees of freedom in the chi-square test.

**eTable 4: ACT-DL versus CBTp exploratory analysis.**

#### Differences in CAARMS distress scores between ACT-DL+TAU and TAU only versus ACT-DL+TAU and CBTp+TAU at baseline, post-intervention, 6-month and 12-month follow-up.

| | | | Adjusted mean difference[a] | | | | Contrasts[b] | | | |
|---|---|---|---|---|---|---|---|---|---|---|
| | Mean (SD), No. | Mean (SD), No. | Mean (SE) | 95% CI | P value | | $\chi^2(3)$ | P value | No. | P value |
| **Time** | **ACT-DL+TAU** | **TAU only** | | | | **Time** | | | | |
| Base | 205·57 (82·84), 70 | 184·88 (78·60), 48 | | | | Base | | | | |
| Post | 76·08 (79·51), 50 | 83·63 (86·12), 35 | 3.39 (16.39) | -28·72 to 35·51 | 0.84 | Post | 1.26 | 0.74 | 115 | 0.70 |
| FU6 | 77·68 (76·23), 41 | 59·6 (69·33), 25 | -17.35 (18.70) | -54·01 to 19·31 | 0.35 | FU6 | | | | 0.44 |
| FU12 | 64·34 (78·57), 41 | 59·6 (60·34), 25 | -0.22 (20.56) | -40·51 to 40·07 | 0.99 | FU12 | | | | 0.27 |
| **Time** | **ACT-DL + TAU** | **CBTp + TAU** | | | | | | | | |
| Base | 205·57 (82·84), 70 | 230·37 (86·61), 27 | | | | | | | | |
| Post | 76·08 (79·51), 50 | 97·17 (88·38), 23 | 11·65 (19·45) | -26·46 to 49·77 | 0.55 | | | | | |
| FU6 | 77·68 (76·23), 41 | 82·71 (96·56), 17 | 1·51 (21·72) | -41·05 to 44·08 | 0.94 | | | | | |
| FU12 | 64·34 (78·57), 41 | 103·13 (100·16), 16 | 29·75 (24·20) | -17·68 to 77·19 | 0.22 | | | | | |

Abbreviations: Base = baseline; Post = post-intervention; FU6 = 6 month follow-up; FU12 = 12 month follow-up; SD = standard deviation; No. = number; SE = standard error; CI = confidence interval.

[a] Reflects the adjusted mean difference between ACT-DL+TAU and TAU only (top) and the adjusted mean difference between ACT-DL+TAU and CBTp+TAU (bottom).

[b] Tests whether contrast 1 (ACT-DL+TAU vs TAU only) is > than contrast 2 (ACT-DL + TAU vs CBTp + TAU) at all three time points and per time point separately.
